# NEURONAL AND ASTROCYTIC TETRAPLOIDY IS INCREASED IN DRUG-RESISTANT EPILEPSY

**DOI:** 10.1101/2022.02.26.22271539

**Authors:** Ancor Sanz-García, Patricia Sánchez-Jiménez, Inmaculada Granero-Cremades, María De Toledo, Paloma Pulido, Marta Navas, Jose Maria Frade, Desirée Pereboom, Cristina Virginia Torres-Díaz, María Carmen Ovejero-Benito

## Abstract

**Background:** Epilepsy is one of the most prevalent neurological diseases, a third of patients remain drug-resistant. The exact etiology of drug-resistant epilepsy (DRE) is still unknown. Neural tetraploidy has been associated with neuropathology. The aim of this study was to assess the presence of tetraploid neurons and astrocytes in DRE.

**Materials & methods:** Cortex, hippocampus and amygdala samples were obtained from patients subjected to surgical resection of the epileptogenic zone. Postmortem brain tissue of subjects without previous records of neurological, neurodegenerative or psychiatric diseases were used as controls. The percentage of tetraploid cells was measured by immunostaining of neurons (NeuN) or astrocytes (S100β) followed by flow cytometry analysis. Results were confirmed by image cytometry (ImageStream X Amnis System Cytometer) and with an alternative astrocyte biomarker (NDRG2). Statistical comparison was performed using univariate tests.

**Results:** A total of 22 patients and 10 controls were included. Tetraploid neurons and astrocytes were found both in healthy individuals and DRE patients in the three brain areas analyzed: cortex, hippocampus and amygdala. DRE patients presented a higher number of tetraploid neurons (p=0.020) and astrocytes (p=0.002) in the hippocampus compared to controls. These results were validated by image cytometry.

**Conclusion:** We demonstrated the presence of both tetraploid neurons and astrocytes in healthy subjects and increased levels of both cell types in DRE patients. This is the first time that tetraploid astrocytes are described in healthy subjects. Furthermore, these results provide new insights into epilepsy, opening new avenues for future treatment.

## INTRODUCTION

Epilepsy is one of the main neurological diseases and affects over 50 million people worldwide. Despite the continuing emergence of new anticonvulsant drugs (AEDs), the percentage of drug-resistant patients is difficult to reduce and remains approximately 25-30% of epileptic patients [1]. The International League Against Epilepsy (ILAE) defined Drug-Resistant Epilepsy (DRE) as a failure of adequate trials of two tolerated, appropriately chosen, and used AED schedules (in monotherapies or in combination) to achieve sustained seizure freedom [2]. Several mechanisms have been hypothesized to explain the processes occurring in the epileptogenic zone of DRE patients: disease-related mechanisms, drug-related mechanisms, and genetic mechanisms, which may be interlinked [3].

Temporal Lobe Epilepsy (TLE) is one of the main types of epilepsy associated with DRE [4]. TLE usually progress with hippocampal sclerosis (HS) that involves massive neuronal loss, especially in the hilar and CA1 (cornus ammonis1) and CA3 regions [5] although it is not completely clear whether this loss is a cause or a consequence [6]. Besides, HS is associated with gliosis, granule cell dispersion, and mossy fiber sprouting [1,7,8].

Tetraploid neurons (with a DNA content equivalent to 4C) can be found in different regions of the healthy central nervous system, such as the cortex and hippocampus [9,10]. These neurons present larger dendritic trees and somas than diploid neurons and contribute to cell variability, playing specific roles as long-distance projection neurons [9–12]. The process causing tetraploidy, cell cycle re-activation, has been observed in different neurodegenerative diseases such as Alzheimer disease [13]. In fact, polyploid neurons are more prone to die [14–16] probably by progressing through the G2/M checkpoint [12,13,17]. This suggests that in neurodegenerative diseases, where neurons become tetraploid, G2/M transition blockade may act as a survival mechanism for the affected neurons [13].

TLE is characterized by recurrent focal seizures originating from a network located discretely along the mesial aspect of the temporal lobe, which are usually accompanied by temporal or HS [4]. So far, HS is the main prognosis marker for surgical resection of focal DRE [8]. In fact, neuronal death in the epileptogenic zone associated with HS occurs in conjunction with electrophysiological changes [1]. Thus, we hypothesized that tetraploid neurons are involved in epilepsy and, more precisely, in HS. These neurons may be more prone to die similarly to tetraploid neurons in Alzheimer patients [14,15,18]. Moreover, we hypothesized that astrocytes could also become tetraploid and contribute to the gliosis observed in HS.

Thus, the first objective was to determine whether tetraploid neurons are involved in TLE. Secondarily, we intended to analyze whether tetraploid astrocytes are present in the brain of DRE patients and control subjects and if they are also involved in TLE.

## MATERIALS AND METHODS

### Study subjects

The protocol and the Informed Consent Form were approved by the Independent Clinical Research Ethics Committee of the Hospital Universitario de La Princesa. The study followed the STROBE guidelines and the Revised Declaration of Helsinki. The epileptogenic zone and the surrounding cortical area were collected from DRE patients and frozen in dry ice immediately after neurosurgical extraction. If the resection involved the amygdala, this tissue was also analyzed. An extensive analysis of patients’ clinical records was performed. Inclusion criteria were patients subjected to resective neurosurgery of the epileptogenic zone who signed the informed consent. The recruitment period lasted 2 years starting on September 2018.

Postmortem tissue of subjects without previous records of neurological, neurodegenerative or psychiatric diseases, who had signed informed consents, was provided by the biobank “Biobanco en Red de la Región de Murcia”, BIOBANC-MUR. Control samples were processed following standard operating procedures with appropriate approval of the Ethics and Scientific Committees.

### Cell nuclei isolation and immunostaining

A 4-5 mm edge cube of human brain tissue was placed in a Dounce homogenizer containing 3.0ml ice-cold, DNase-free PBS 0.1%Triton (PBT) with a protease inhibitor cocktail (Roche, Switzerland). Cell nuclei isolation was carried out following a modified form of a previously reported procedure [10,19].

Immunostaining was performed by adding primary and secondary antibodies simultaneously to isolated unfixed nuclei containing 5% fetal calf serum (FCS) and 1.25mg/ml BSA. In control samples, the primary antibodies were excluded. Finally, the reaction was incubated O/N at 4°C in the dark. Immunostained nuclei were filtered through a 40μm nylon filter, able to retain big aggregates but not nuclei. Then, the volume was adjusted to 350-800μl with DNase-free PTx containing 40μg/ml propidium iodide (PI, Sigma, USA) and 25μg/ml DNAse-free RNAse I (Sigma, USA), and analyzed in a Flow cytometer or an ImageStream X Amnis System Cytometer equipment.

The rabbit anti-NeuN polyclonal antibody (Merck, Germany) was diluted 1/800 and detected with a donkey anti-rabbit Alexa Fluor 488 (Invitrogen, UK). The mouse S100β monoclonal antibody [4C4.9] (Thermo Fisher Scientific, USA), was used at 1/70 dilution and detected with a goat anti-rabbit Alexa Fluor 647 (Invitrogen, USA).

The rat anti-CTIP2 monoclonal antibody [25B6] (Abcam, UK) was used at 6µg/ml. The rabbit polyclonal Anti-NDRG2 Antibody HPA002896 (Atlas antibodies) was used at 4µg/ml. These antibodies were detected with a goat anti-rabbit DyLight 405 (Invitrogen, USA) and an anti-rat 647 (Atlas antibodies, Sweden) and used at 1/500.

### Flow cytometry

NeuN and S100β analysis were carried out in the Flow Cytometry Unit of Hospital de La Princesa using a FACSCanto II Cytometer (BD Biosciences, USA). NDRG2 and CTIP2 analysis were carried out in an Attune™ NxT Acoustic Focusing Cytometer (Thermo Fisher Scientific Inc., USA) in the Flow Cytometry Unit of “Instituto de Medicina Molecular Aplicada” (IMMA, Universidad San Pablo CEU) (Figure 1).

**Figure 1:**
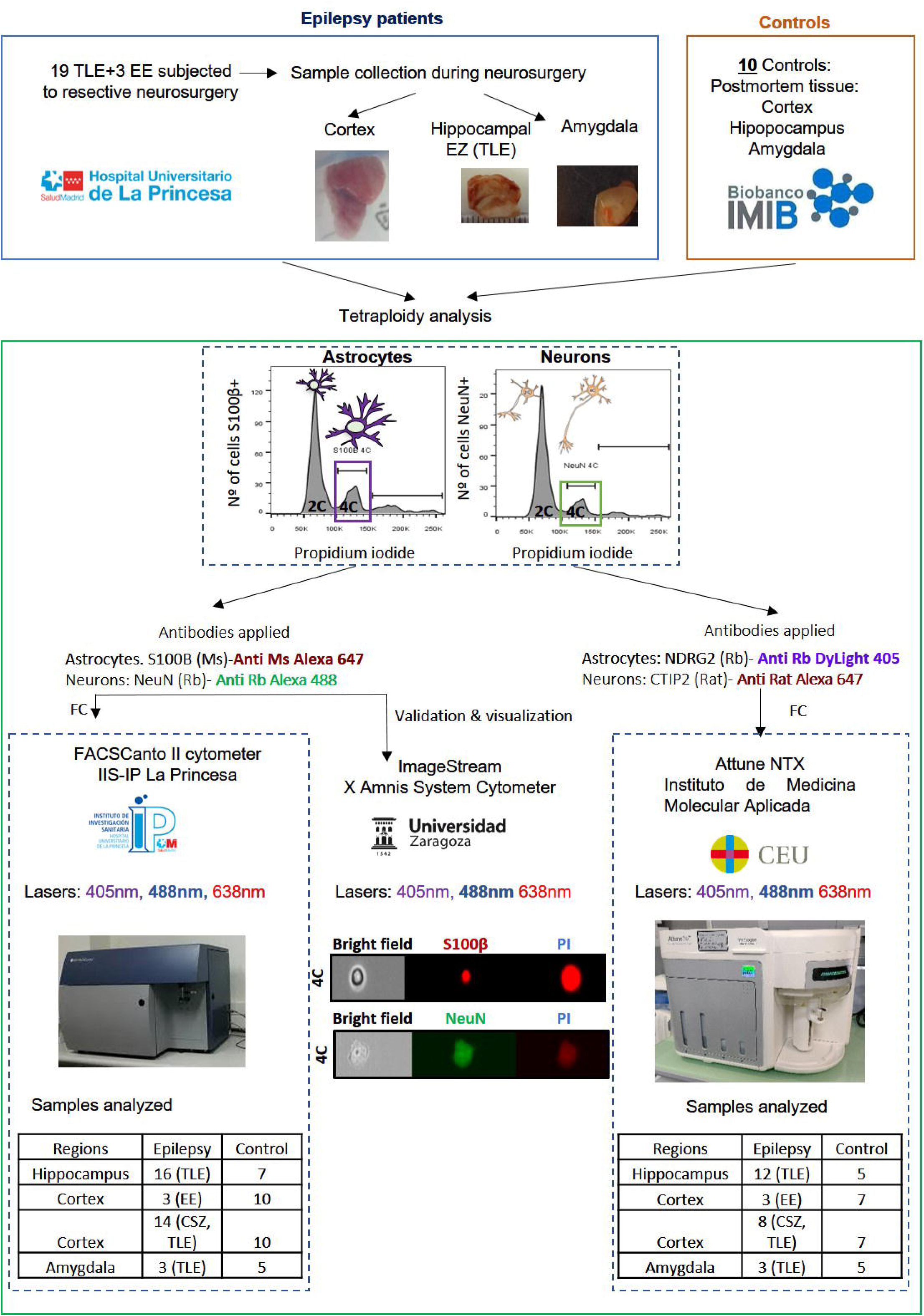
Workflow of the experiments performed, the samples analyzed and the equipment used for each determination. Abbreviations: 2C: diploid neurons 4C: tetraploid neurons, CSZ: cortical surrounding zone to the epileptogenic zone; EE: extratemporal epilepsy; EZ epileptogenic zone; Ms: mouse; PI: propidium iodide, RB: rabbit; TLE: temporal lobe epilepsy.

Data were analyzed with FACSDiva (BD Biosciences), Thermo Scientific™ Attune™ NxT Software, (Thermo Fisher Scientific, USA) and FlowJo (Walter and Eliza Hall Institute of Medical Research, Australia) and displayed using logarithmic scaling. Tetraploidy was analyzed as described previously focusing on the percentage of tetraploid neurons or astrocytes [10] (Supplementary Figure 1).

### Tetraploid cells visualization

An ImageStreamX Amnis System Cytometer (EMD Millipore, USA) that combine confocal microscopy and flow cytometry was used to visualize tetraploid neurons and astrocytes immunostained with NeuN and S100β as described above (Supplementary Figure 2).

### Statistical analysis

Quantitative variables are expressed as mean and standard deviation. Statistical comparison was performed by the Wilcoxon rank-sum test. The resulting comparison was represented by using boxplots combined with density plots, which allow representing the patients’ distribution according to the percentage of tetraploidy (wider densities imply a higher number of patients for that percentage). For clarity, the density plot is not shown for groups with low dispersion. The relation between clinical data and tetraploidy results was assessed by using the Pearson correlation coefficient. p<0.05 was considered statistically significant. Statistical analyses were performed in R, version 4.0.3 (http://www.R-project.org).

## RESULTS

### Study population

Twenty-two DRE patients subjected to neurosurgical resection of the epileptogenic zone and 10 controls were recruited (Table 1). Nineteen patients were diagnosed with TLE, with and epileptogenic zone located in the hippocampus, 3 patients had extratemporal epilepsies. Most of the patients (81%) achieved Engel I/II [20] 6 months after neurosurgery and thus were considered responders (Table 1).

**Table 1:** Clinical and sociodemographic data of DRE patients and controls included in this study.

|  |  | Patients | Controls |
| --- | --- | --- | --- |
|  | Women (%) | 9 (39%) | 3 (30%) |
|  | Age (years) | 46 ± 9 | 60 ± 12 |
|  | Age at onset (years) | 18 ± 14 | NA |
|  | Seizures/month (n) | 7 ± 8 |  |
|  | Anticonvulsant drugs (n) | 5 ± 3 |  |
| Seizure etiology | Structural | 5 (23%) |  |
|  | Infectious | 2 (9%) |  |
|  | Unknown | 15 (68%) |  |
| % Engel I/II | 6 months | 17 (81%)<sup>#</sup> |  |
|  | 12 months | 14 (88%)<sup>#</sup> |  |
|  | 24 months | 11 (85%)<sup>#</sup> |  |

Data are presented as mean ± standard deviation or number (%). Outcome of neurosurgical resection was evaluated using Engel's classification at 6, 12 and 24 months after surgery. Patients were classified as responders to neurosurgery if they achieved Engel I and II. # Response was calculated for those patients whose clinical information was available. Abbreviations: NA, not applicable.

### Neuronal tetraploidy

Initially, the percentage of tetraploid neurons present in the epileptogenic zone was estimated using flow cytometry as the proportion of positive cells for the neuronal marker NeuN [10,19]. The number of cytometry determinations varied among the different experimental conditions due to experimental issues (see Methods, Figure 1). Only mature neurons that expressed NeuN in absence of S100β were considered. TLE patients (n=16) showed a significant increase in the percentage of tetraploid neurons in the epileptogenic zone of the hippocampus with respect to controls (n=7, p=0.020, Figure 2A). As can be observed in the density plot (Figure 2A), in the control group most subjects exhibited a percentage of tetraploid neurons lower than 1, which was increased to 1.5 in the epilepsy group. To determine if the increase of neuronal tetraploidy was specific to the hippocampus, the percentage of tetraploid neurons was also analyzed in extratemporal epilepsies (EE). An increase in the percentage of tetraploid neurons was observed in the cortical epileptogenic zone (n=3) compared with the control cortex (n=10). Although this difference was not significant, its p-value was very close to the significance threshold (p=0.052, Figure 2B). The percentage of tetraploidy in the amygdala could be assessed in patients who were subjected to amygdalohippocampectomy. No significant differences were observed between the amygdala of controls (n=3) and patients (n=5, p=0.229, Figure 2D). Finally, to determine if this increase in neuronal tetraploidy was specific for the epileptogenic zone, the surrounding region to the epileptogenic zone that needed to be removed to access the hippocampus was also analyzed. No significant differences were observed between the surrounding cortical region of epileptic patients (n=14) and control subjects (n=10, Figure 2C). Similarly, the comparison between the epileptogenic cortical region (n=3) of those patients with extratemporal epilepsy and the surrounding cortical zone of TLE patients (n=14) showed no significant difference (p=0.438, Supplementary Figure 3A). Results of the tetraploidy analysis in double-positive (NeuN+ and S100β+) cells can be observed in Supplementary Figure 3.

**Figure 2.**
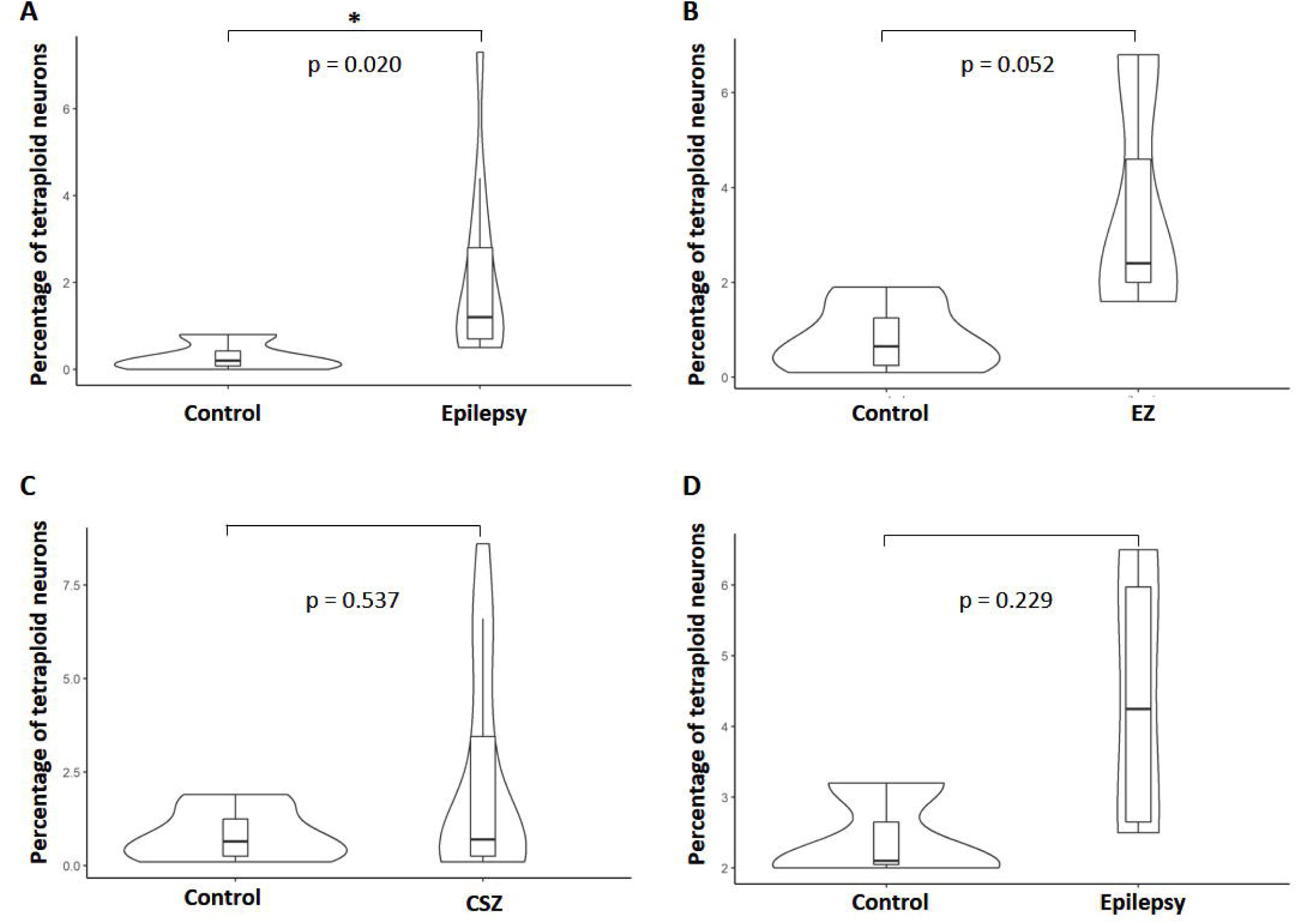
Percentage of tetraploid neurons in different brain regions of TLE patients and controls. Cells from surgical samples were analyzed for marker expression and tetraploidy by flow cytometry. Percentage of tetraploid neurons (NeuN+ S100β-cells) is shown for: A) Hippocampus of healthy controls (n=7) and the epileptogenic zone of TLE patients (n=16). B) Cortex of healthy controls (n=10) and epileptogenic zone of extratemporal epilepsy (EZ, n=3). C) Cortex of healthy controls (n=10) and cortical surrounding zone (CSZ, n=14) to the epileptogenic zone. D) Amygdala of healthy controls (n=5) and of TLE patients (n=3). Data are represented as violin plots. Differences were analyzed by Wilcoxon rank sum exact test. * p<0.05. Abbreviations: CSZ: cortical surrounding zone to the epileptogenic zone; EZ epileptogenic zone.

Moreover, to confirm the presence of tetraploid neurons in both patients and controls, tetraploid neurons were visualized using AMNIS FlowSight equipment. This analysis indicated that tetraploid neurons from both controls and patients showed a rounded morphology with just one nucleus with double DNA content (Figure 3A).

**Figure 3.**
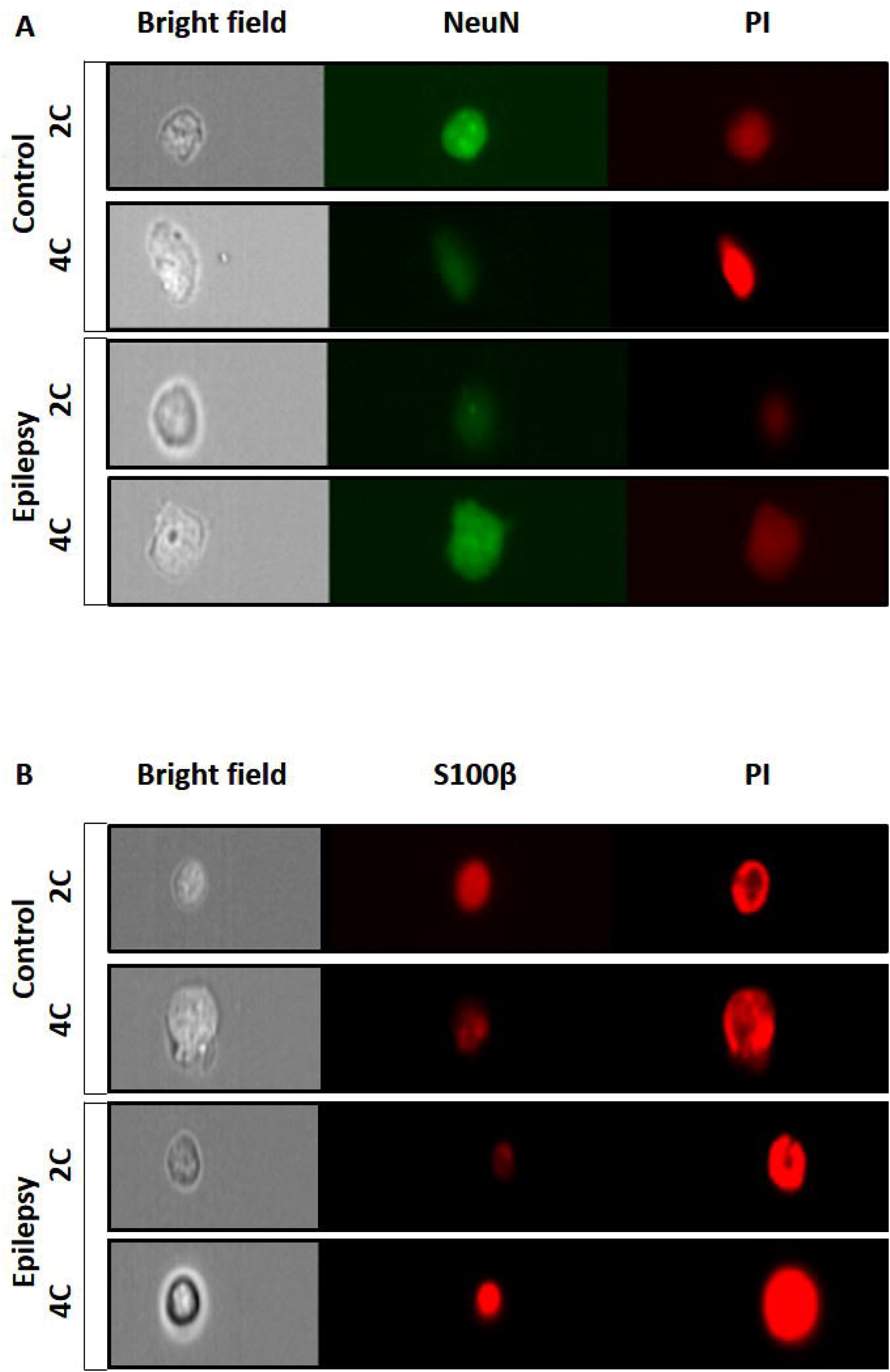
Visualization of tetraploid neurons from TLE patients and controls. A) Cells from surgical samples were stained for NeuN and with PI, and images were acquired and analyzed with an ImageStream X Amnis System Cytometer. Bright field images (left panels) and NeuN (middle panels) and PI (right panels) staining are shown for NeuN+ cells from healthy control subjects (control) and TLE patients (epilepsy). 3B). Cells from surgical samples were stained for S100β and with propidium PI, and images were acquired and analyzed with an ImageStream X Amnis System Cytometer. Bright field images (left panels) and S100β (middle panels) and PI (right panels) staining are shown for S100β+ cells from healthy control subjects (control) and TLE patients (epilepsy). Abbreviations: 2C: diploid cells 4C: tetraploid cells, PI: propidium iodide.

The presence of tetraploid neurons was also analyzed in a subset of patients using a different biomarker of differentiated neurons (CTIP2), which has been shown to be enriched in the tetraploid population. Overall, no differences were found between controls and DRE patients in any of the regions analyzed (hippocampus and cortex) (Supplementary Figure 4).

### Astrocytic tetraploidy

Although neuronal dystrophy is one of the main hallmarks of HS, glia plays a key role in the epileptogenic zone. Thus, we wondered if tetraploid astrocytes could be found in DRE patients.

S100β stained tetraploid astrocytes were found in the hippocampus, cortex and amygdala of healthy controls (Figures 3B and 4). Moreover, when the epileptogenic zone was the hippocampus (n=16), levels of tetraploid astrocytes stained with S100β were significantly increased compared with those of the control hippocampus (p=0.002, n=7, Figure 4A). No differences were found in the cortical epileptogenic zone (p=0.926, Figure 4B) nor in the amygdala (p=0.114, Figure 4D). Although the mean of tetraploid astrocytes was increased in the cortex surrounding the epileptogenic zone, this difference was not statistically significant (p=0.144, Figure 4C). No significant differences were observed between the epileptogenic zone and the surrounding zone (Supplementary Figure 5B).

**Figure 4.**
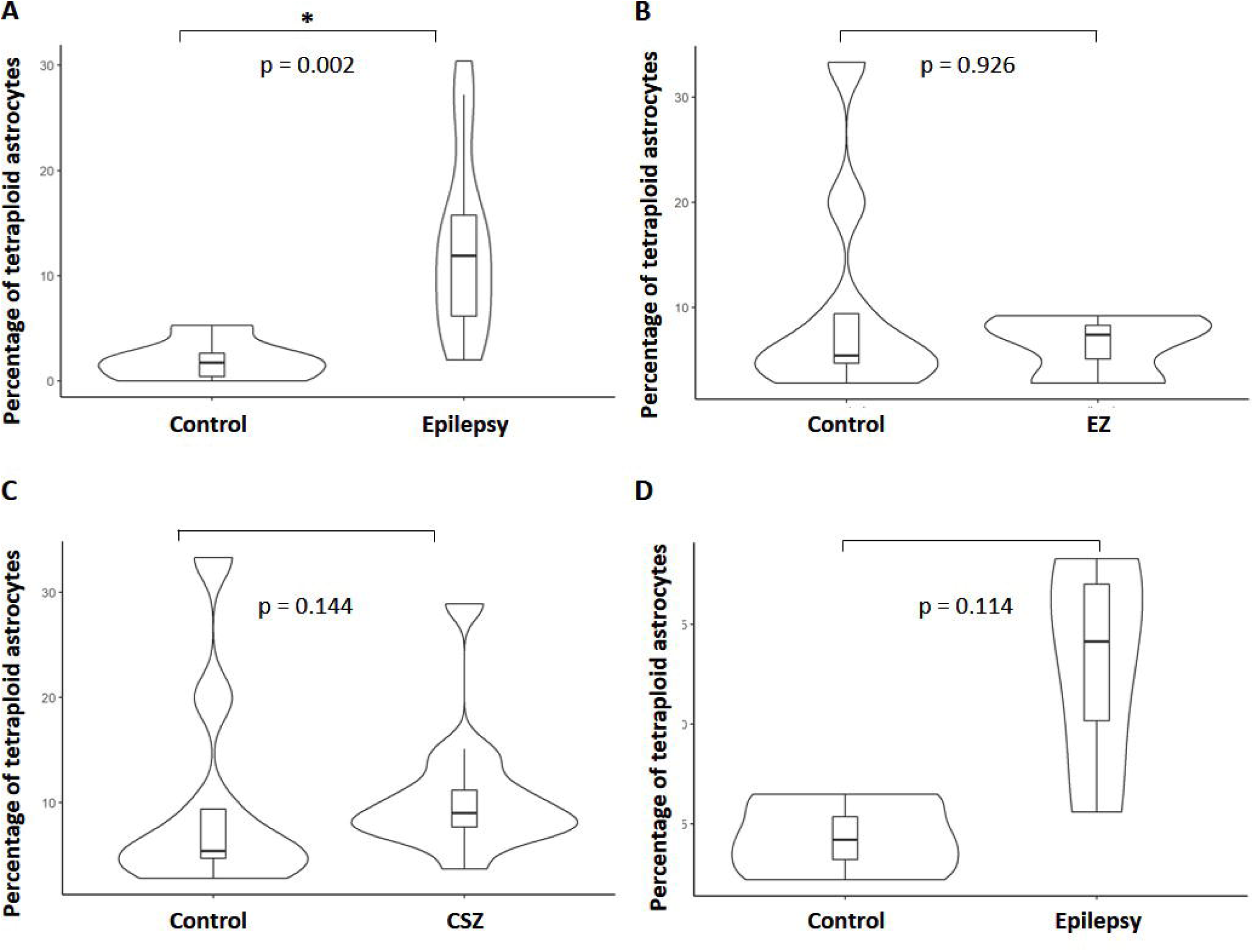
Percentage of tetraploid astrocytes in different brain regions of TLE patients and controls. Cells from surgical samples were analyzed for marker expression and tetraploidy by flow cytometry. Percentage of tetraploid astrocytes (S100β+NeuN-cells) is shown for: A) Hippocampus of healthy controls (n=7) and the epileptogenic zone of TLE patients (n=16). B) Cortex of healthy controls (n=10) and epileptogenic zone of extratemporal epilepsy (EZ, n=3). C) Cortex of healthy controls (n=10) and cortical surrounding zone (CSZ, n=14) to the epileptogenic zone. D) Amygdala of healthy controls (n=5) and of TLE patients (n=3). Data are represented as violin plots. Differences were analyzed by Wilcoxon rank sum exact test. * p<0.05. Abbreviations: CSZ: cortical surrounding zone to the epileptogenic zone; EZ epileptogenic zone.

S100β stained tetraploid astrocytes from the brain of epileptic patients and controls were visualized using an AMNIS FlowSight to confirm cytometry results. These astrocytes showed rounded morphology and only one nucleus (Figure 3B).

An alternative biomarker of differentiated astrocytes (NDRG2) was also used for tetraploidy analysis (Supplementary Figure 6). The percentage of tetraploid astrocytes was higher in the epileptogenic zone of epilepsy patients (13.70±5.48, n=12) compared to the control hippocampus (7.42±5.10, n=5), with a p-value very close to the significance threshold (p=0.051). There were no significant differences between tetraploid astrocytes in the cortical epileptogenic zone and those of the control cortical tissue (p=0.905, Supplementary Figure 6B). Moreover, the pattern of tetraploid astrocytes in the NDRG2 subset was similar in the control cortex and the surrounding zone, and no differences were observed in the amygdala (Supplementary Figure 6 C-D).

Lastly, the relation between tetraploidy and clinical data is shown in Supplementary Figure 7. The correlation between the percentage of tetraploid astrocytes and neurons was very strong for the amygdala (r=0.83), cortex (r=0.98), and hippocampus (r=0.99). Surprisingly, there was a high correlation between the number of anti-convulsant drugs used by the patients and tetraploidy levels in the amygdala (r=0.83 for astrocytes and r=1 for neurons) but not in the other two areas. As expected, there was a high correlation between age and tetraploidy in the cortex (r=0.88 for astrocytes and r=0.95 for neurons). An inverse correlation with age (r=-0.99 for astrocytes and r=-0.86 for neurons) was found in the amygdala, while no correlation was found in the hippocampus. Interestingly, there was a high inverse correlation between onset age and tetraploidy (r=-0.95 for astrocytes and r=-0.97 for neurons) suggesting that early onsets implied higher percentages of tetraploidy. In the other two areas no correlation with age of onset was found.

## DISCUSSION

In this study, we demonstrated the presence of both tetraploid neurons and astrocytes in TLE patients and control subjects. We also found a higher number of tetraploid neurons and astrocytes in the hippocampus of TLE patients.

As expected, tetraploid neurons were located in the cortex, hippocampus [10,19] and amygdala of healthy controls. A significant increase in tetraploid neurons detected with NeuN was observed in TLE. We perceived a non-significant increment of tetraploid neurons in the surrounding region to the epileptogenic zone. The increase of extratemporal tetraploidy was very close to the significance threshold (p=0.051), probably due to the limited number of samples available (n=3). To analyze tetraploidy, adult neurons were identified using the NeuN biomarker to differentiate them from adult neurogenesis, which is also impaired and abnormal in TLE [21,22]. Unfortunately, we could not confirm the increase of neuronal tetraploidy in the hippocampus of TLE patients with an alternative biomarker (CTIP2). Although CTIP2 levels in tetraploid neurons were higher than those reported in previous publications, this marker is only expressed in a subset of tetraploid neurons [10].

Cell cycle reactivation in neurons from epilepsy patients has been proposed to modulate apoptosis [23–25]. Several factors were described in this process, such as E2F1, implicated in neuronal cell cycle reactivation and DNA synthesis [12], which is found in an abnormal location in the neuronal cytoplasm of neurons of TLE patients [26]. Cyclin B, involved in G2/M transition, has been observed in the hippocampus of TLE patients [25]. Indeed, BDNF, a blocker of G2/M transition by inhibiting Cdk1 in physiological tetraploid neurons [17], is overexpressed during seizures [27,28]. These tetraploid neurons may be more prone to die, as previously described in Alzheimer disease [14,15,18], contributing to the massive apoptotic cell death observed in the hippocampus of TLE patients [26,29]. Nevertheless, we could not analyze the association between neuron tetraploidy and cell death.

Most of the DRE studies have focused on neurons [30]. However, astrocytes modulate synaptic plasticity and electrical properties in the brain in physiological conditions [31– 34]. Recent studies have revealed that reactive astrocytes actively contribute to seizure activity in epileptogenesis [30]. During this process, astrocytes undergo morphological, molecular, and functional modifications that allow the development of astrogliosis, along with changes in the expression pattern of various proteins, such as transporters, receptors, and enzymes, some of which are involved in astrocyte–neuron and astrocyte-astrocyte signaling [30]. In the 60s, there was controversy regarding the existence of tetraploid astrocytes and neurons [35–38]. As there were no reproducible methods of DNA quantification, different researchers stated that it was impossible to reach a conclusion about this topic [39]. We have observed the presence of tetraploid astrocytes in different regions (cortex, hippocampus and amygdala) of the healthy central nervous system and TLE with two different biomarkers (S100β and NDRG2) and two techniques (Flow Cytometry and AMNIS FlowSight). We have confirmed that tetraploid astrocytes are increased in TLE. Tetraploid astrocytes may be bigger than diploid astrocytes, thereby contributing to the hypertrophy/hyperplasia of astrocytes found in the epileptogenic zone of TLE [40,41]. Recently, polyploid astrocytes with abnormal mitosis were described in animal models of epilepsy [42] and Drosophila [43]. These studies found that multinucleated astrocytes could survive long periods and even re-enter mitosis [42]. These results suggest that possible complementary mechanisms caused by insults generated by seizures could alter the cell cycle.

Different processes have been observed in TLE patients [40,41,44], suggesting cell cycle reactivation and proliferation in astrocytes. Surprisingly and contrary to the processes observed in neurons, NGF and p75 promote cell cycle arrest in astrocytes in epilepsy [45]. Moreover, in mouse models of TLE, BDNF and TrkB regulate severity and neuronal activity [46], which may contribute to the G2/M blockade in tetraploid astrocytes in a similar way to the modulation of tetraploid neurons [17].

In accordance with previous results [10], we found, in neurons and astrocytes, a high correlation between age and cortical tetraploidy and between lower age of onset and high levels of tetraploidy. Thus, if patients show an early onset of epilepsy, they may accumulate higher levels of tetraploidy. Moreover, an association between tetraploid neurons and tetraploid astrocytes was detected. This fact may be potentially explained if epilepsy could stimulate overexpression of factors that induce cell cycle re-entry (such as neurotrophins) [12,19,47] and tetraploidy in both cell populations. Similarly to recent treatments proposed for Alzheimer [48], we hypothesize that the modulation of tetraploidy could be a potential treatment for DRE.

## Limitations

This work has several limitations. The first and most relevant is the sample size. The number of patients, mainly DRE, is limited by the number of neurosurgical resections. This limitation is balanced by the exhaustive study of the patient clinical records and the fact that the tissue sample was fresh thus precluding potential biases caused by fixation in formalin. Secondly, the control sample came from a brain tissue biobank; therefore, possible effect of death on postmortem tissue cannot be ruled out. Taking all together, further studies are needed to confirm our results. Thirdly, although astrocytes are the brain cell type that expresses the highest S100β [49], we observed that a subpopulation of neurons also expressed this marker [50]. Thus, we had to focus on S100β+/NeuN-astrocytes. Fourthly, the present study does not rule out the possible increase of tetraploid neurons and astrocytes in drug-sensitive patients; the obvious limitation of tissue samples from drug-sensitive patients hinders the comparison between drug-sensitive and DRE.

## Conclusions

The presence of both tetraploid neurons and astrocytes in healthy subjects is a groundbreaking discovery that could provide new insights into the study of the brain. The role of these cells in the structure and function of the brain remains unexplored and should be better studied in the future. Furthermore, these cells are increased in DRE. Accordingly, understanding the role of these cells in the pathogenesis of this disease could pave the way to new treatment lines for these patients.

## Data Availability

All data produced in the present study are available upon reasonable request to the authors.

## Acknowledgments

We thank to the epilepsy patients who kindly donated brain samples. We are particularly grateful for the generous contribution of the patients and the collaboration of Biobank Network of the Region of Murcia, BIOBANC-MUR, registered on the Registro Nacional de Biobancos with registration number B.0000859. BIOBANC-MUR is supported by the “Instituto de Salud Carlos III” (proyecto PT20/00109), by “Instituto Murciano de Investigación Biosanitaria Virgen de la Arrixaca, IMIB” and by “Consejería de Salud de la Comunidad Autónoma de la Región de Murcia”. We would like to thank Javier Fraga and Manuel Gómez Gutierrez for their help with the study and their valuable comments on this manuscript. We also thank our colleges from the Flow Cytometry Unit of “Instituto de Medicina Molecular Aplicada” (IMMA, Universidad San Pablo CEU).

## Data availability statement

All data produced in the present study are available upon reasonable request to the authors.

## FIGURES AND TABLES

**Supplementary Figure 1**. Gating strategy for assessment of tetraploid cell populations by Flow cytometry. A) Nuclei were gated by forward scattering area (FCS-A), a measure of size, and side scattering area (SSC-A), a measure of complexity B) Nuclei were differentiated from debris by their ability to incorporate propidium iodide. C) DNA content was assessed from the gated nuclear population by plotting Propidium Iodide-H versus Propidium Iodide-A levels. Diploid (2C) and tetraploid (4C) nuclei were found on the diagonal of the plot and the doublets of diploid nuclei were excluded. D) Singlets were further gated by plotting Propidium Iodide-W versus Propidium Iodide-A levels. E, F) Representative dot plots of singlets of the fluorescent minus one controls: E) was stained with anti-S100B, Alexa Fluor 488 and Alexa Fluor 647 and F) was stained with anti-NeuN, Alexa Fluor 48 and Alexa Fluor 647. G) Representative histogram showing DNA content of S100β+ NeuN-cells of cortical tissue from a control subject (pink) and a DRE patient (blue). H) Representative histogram showing DNA content of S100β-NeuN+ cells of cortical tissue from a control subject (pink) and a DRE patient (blue). Abbreviations: 2C: diploid cells 4C: tetraploid cells, PI: propidium iodide.

**Supplementary Figure 2**. Gating strategy for analysis of tetraploid cell populations by ImageStream X Amnis System Cytometer. A) Bi-parametric dot plot representing Aspect Ratio_M01 / Area_M01. Single nuclei were separated from doublets by gating as indicated in the dot plot, followed by a revision of images of all cells. B) Example of the visualization of a clump of cells (1) and a single cell (2) belonging to the gate Singlets. C) NeuN expression on singlets based on the expression of the fluorophore Alexa Fluor 488. The gate used to identify neurons is shown within the histogram. D) DNA content of neurons. The gate used to identify tetraploid neurons (4C) is shown within the histogram. The images of the cells selected with this gate were also checked to confirm the presence of singlets and discard the presence of clumps of nuclei. E) S100β expression on singlets based on the expression of the fluorophore Alexa Fluor 647. The gate used to identify astrocytes is shown within the histogram. F) DNA content of astrocytes. The gate used to identify tetraploid astrocytes (4C) is shown within the histogram. The images of the cells selected with this gate were also checked to confirm the presence of singlets. Abbreviations: 2C: diploid cells 4C: tetraploid cells.

**Supplementary Figure 3**. Percentage of NeuN+S100β+ tetraploid cells in different brain regions. Cells from surgical samples were analyzed for marker expression and tetraploidy by flow cytometry. Percentage of NeuN+S100β+ tetraploid cells is shown for: A) Hippocampus of healthy controls (n=7) and the epileptogenic zone of TLE patients (n=16). B) Cortex of healthy controls (n=10) and epileptogenic zone of extratemporal epilepsy (EZ, n=3). C) Cortex of healthy controls (n=10) and cortical surrounding zone (CSZ, n=14) to the epileptogenic zone. D) Amygdala of healthy controls (n=5) and of TLE patients (n=3). Data are represented as violin plots. Differences were analyzed by Wilcoxon rank sum exact test. * p<0.05. Abbreviations: CSZ: cortical surrounding zone to the epileptogenic zone; EZ epileptogenic zone.

**Supplementary Figure 4**. Percentage of tetraploid neurons assessed with CTIP2 in different brain regions of TLE patients and controls. Cells from surgical samples were analyzed for marker expression and tetraploidy by flow cytometry. Percentage of tetraploid neurons (CTIP2+ cells) is shown for: A) Hippocampus of healthy controls (n=5) and the epileptogenic zone of TLE patients (n=12). B) Cortex of healthy controls (n=7) and epileptogenic zone of extratemporal epilepsy (EZ, n=3). C) Cortex of healthy controls (n=7) and cortical surrounding zone (CSZ, n=8) to the epileptogenic zone. Data are represented as violin plots. Differences were analyzed by Wilcoxon rank sum exact test. * p<0.05. Abbreviations: CSZ: cortical surrounding zone to the epileptogenic zone; EZ epileptogenic zone.

**Supplementary Figure 5**. Percentage of tetraploid astrocytes and neurons in the cortex. Cells from surgical samples were analyzed for marker expression and tetraploidy by flow cytometry. A) Percentage of tetraploid neurons (NeuN+S100β-cells) in the cortex of healthy controls (n=10) and epileptogenic zone of extratemporal epilepsy (EZ, n=3). B) Percentage of tetraploid astrocytes (NeuN-S100β+ cells) in the cortex of healthy controls (n=10) and epileptogenic zone of extratemporal epilepsy (EZ, n=3). C) Percentage of tetraploid cells (NeuN+ and S100β+ cells) in the cortex of healthy controls (n=10) and the epileptogenic zone of extratemporal epilepsy (EZ, n=3). D) Percentage of tetraploid neurons (CITP2+ cells) in the cortex of healthy controls (n=7) and the epileptogenic zone of extratemporal epilepsy (EZ, n=3). B) Percentage of tetraploid astrocytes (NeuN-NDRG2+ cells) in the cortex of healthy controls (n=7) and the epileptogenic zone of extratemporal epilepsy (EZ, n=3). Differences were analyzed by Wilcoxon rank sum exact test. * p<0.05. Abbreviations: CSZ: cortical surrounding zone to the epileptogenic zone; EZ epileptogenic zone.

**Supplementary Figure 6**. Percentage of tetraploid astrocytes assessed with NDRG2 in different brain regions of TLE patients and controls. Cells from surgical samples were analyzed for marker expression and tetraploidy by flow cytometry. Percentage of tetraploid astrocytes (NDRG2+) is shown for: A) Hippocampus of healthy controls (n=5) and the epileptogenic zone of TLE patients (n=12). B) Cortex of healthy controls (n=7) and epileptogenic zone of extratemporal epilepsy (EZ, n=3). C) Cortex of healthy controls (n=7) and cortical surrounding zone (CSZ, n=8) to the epileptogenic zone. D) Amygdala of healthy controls (n=3) and of TLE patients (n=5). Data are represented as violin plots. Differences were analyzed by Wilcoxon rank sum exact test. * p<0.05. Abbreviations: CSZ: cortical surrounding zone to the epileptogenic zone; EZ epileptogenic zone.

**Supplementary Figure 7**. Correlations between percentage of tetraploid cells and clinical data. Correlations were analyzed by Pearson correlation coefficients. Values of coefficients are indicated within squares. Bright red indicates a strong positive correlation, while dark blue represents a strong negative correlation. Abbreviations: % 4C S100β, percentage of tetraploid astrocytes (S100β+ NeuN-); % 4C S100β, percentage of tetraploid neurons (NeuN+ S100β-).

