## Supplementary figures and images for "NEURONAL AND ASTROCYTIC TETRAPLOIDY IS INCREASED IN DRUG-RESISTANT EPILEPSY"

### Supplementary Figure 1

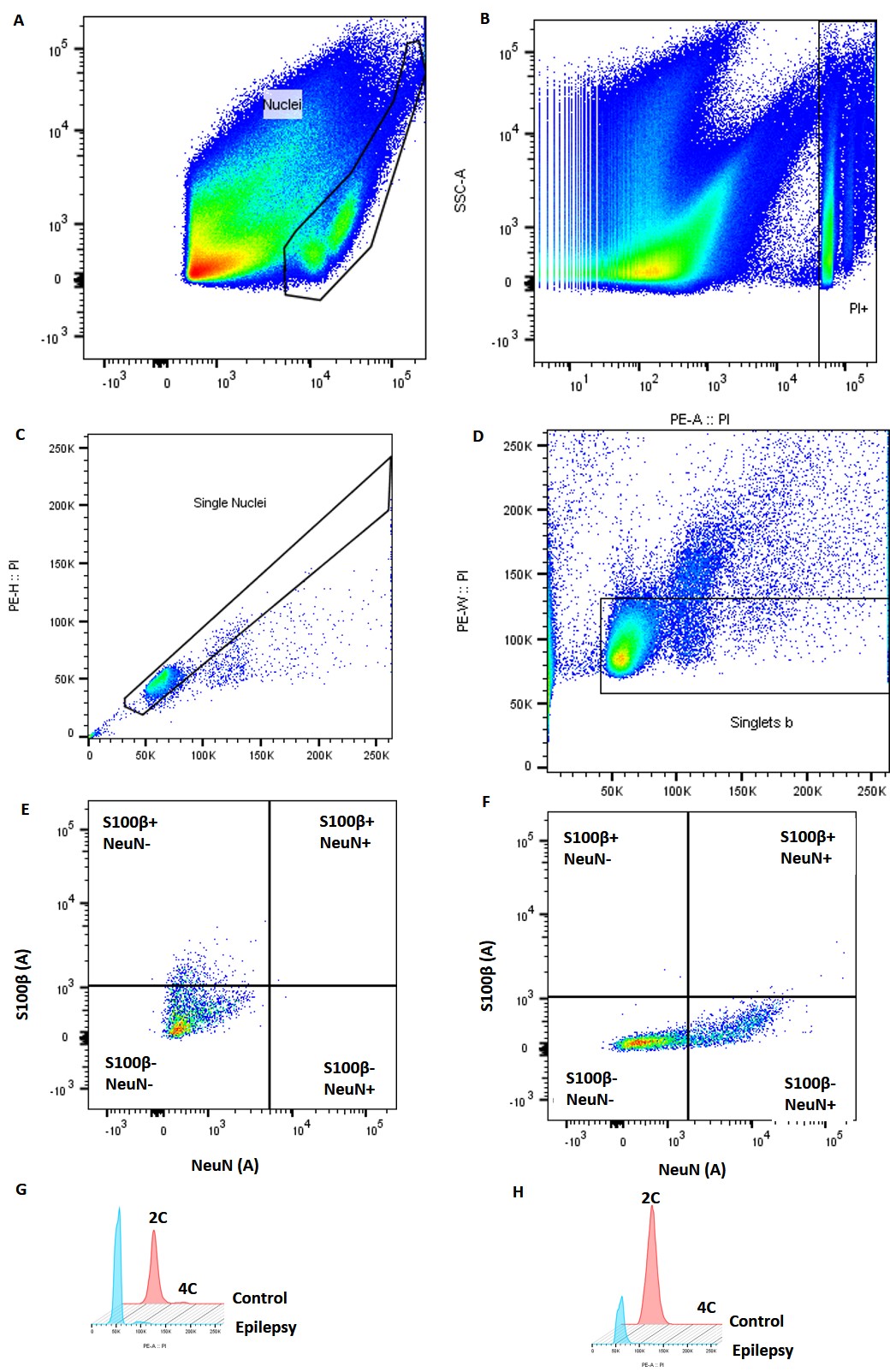

### Supplementary Figure 2

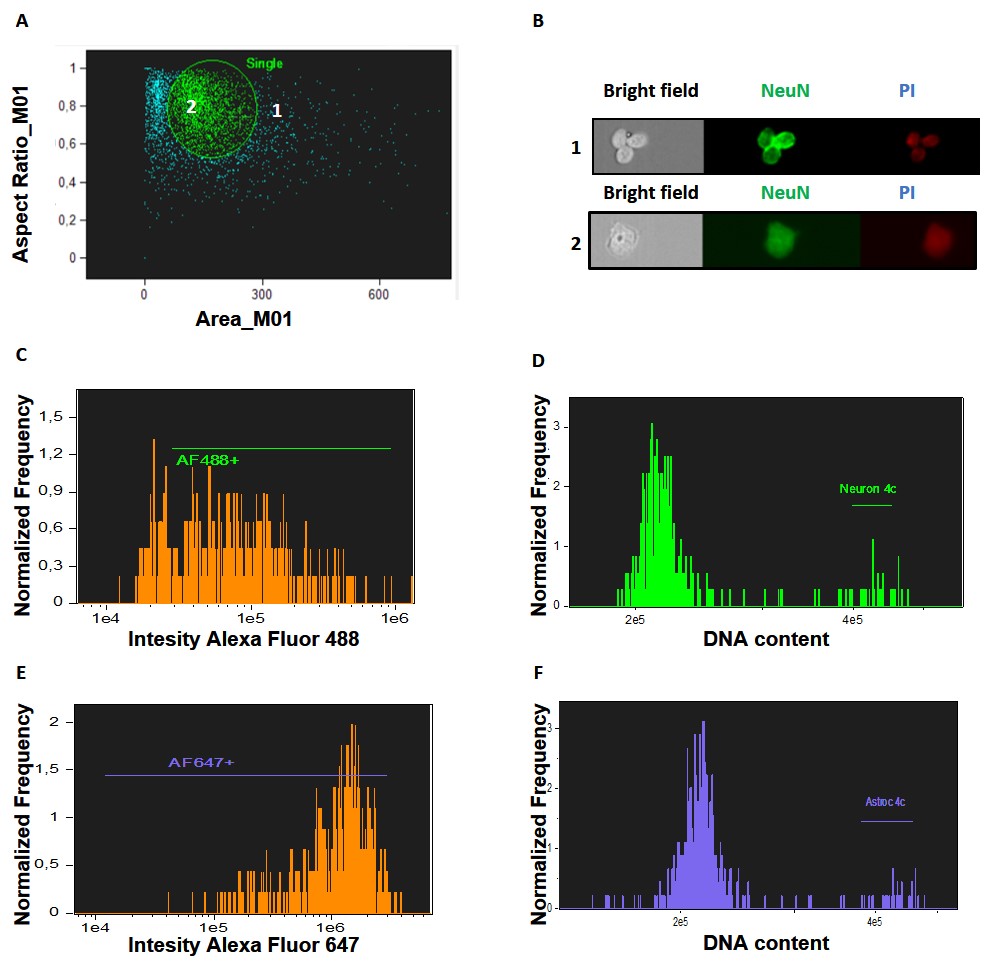

### Supplementary Figure 3

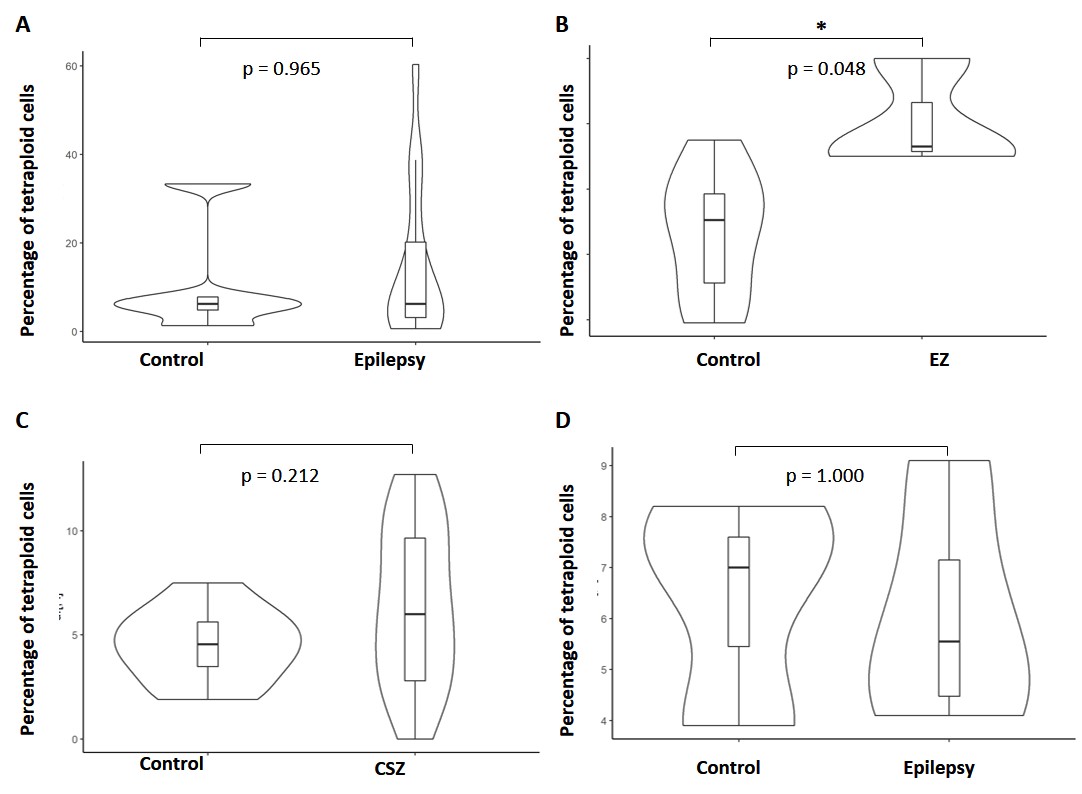

### Supplementary Figure 4

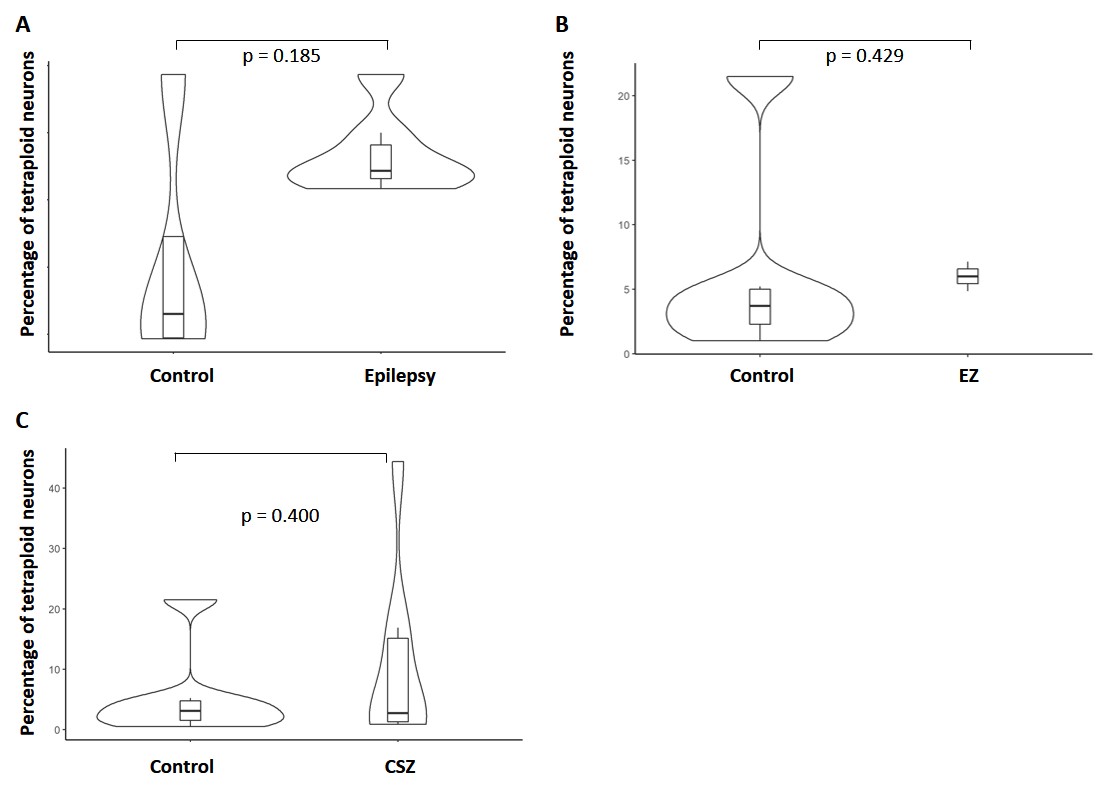

### Supplementary Figure 5

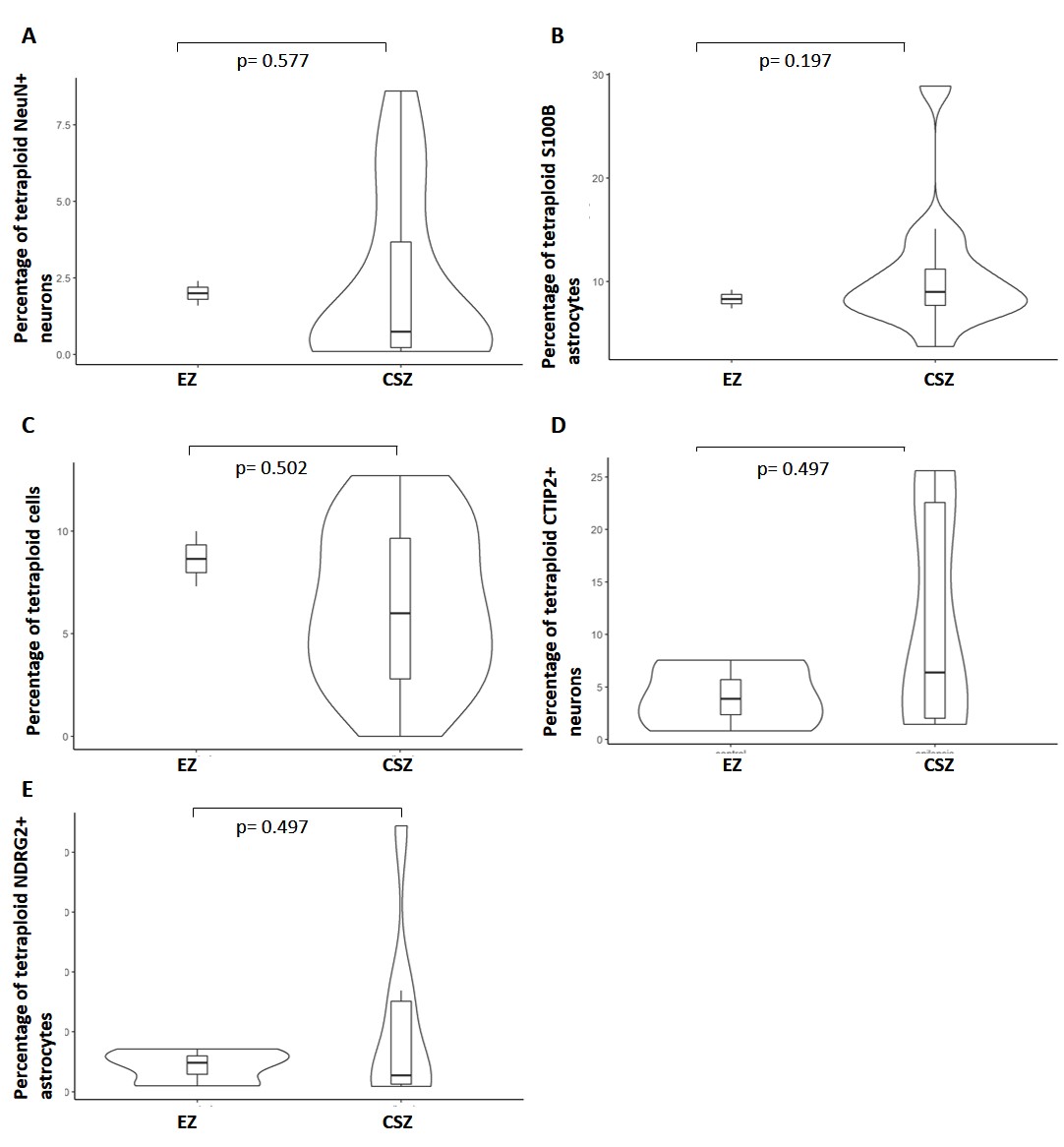

### Supplementary Figure 6

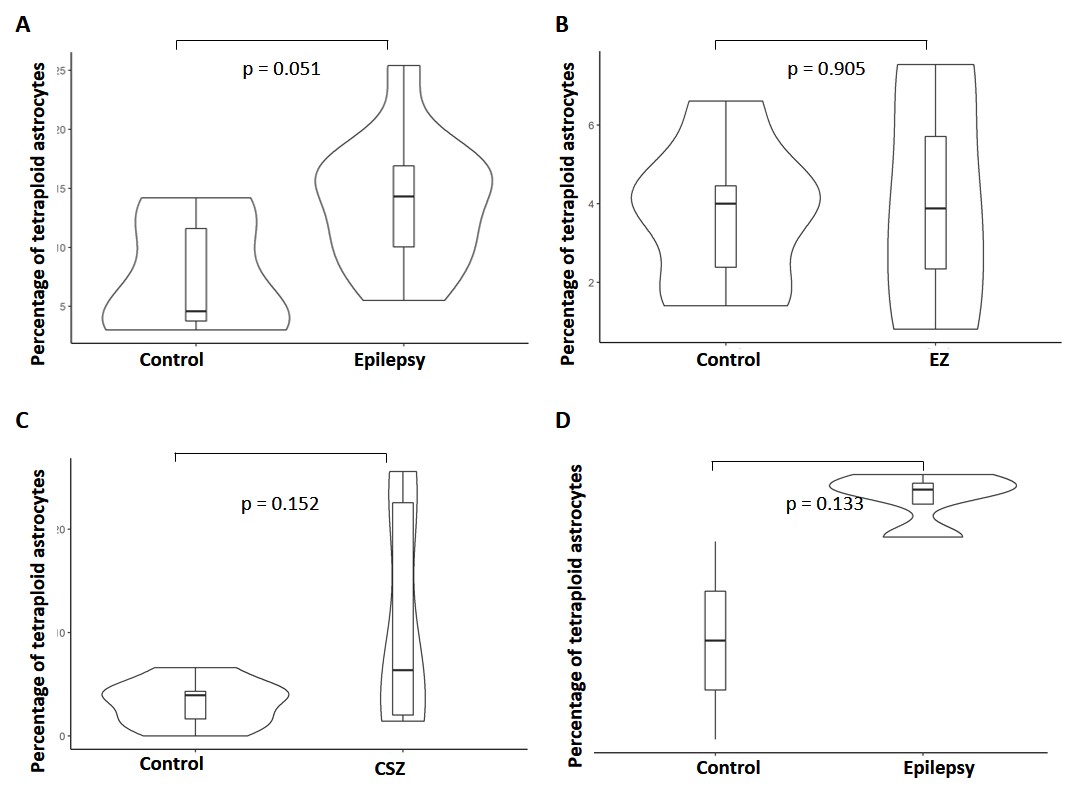

### Supplementary Figure 7

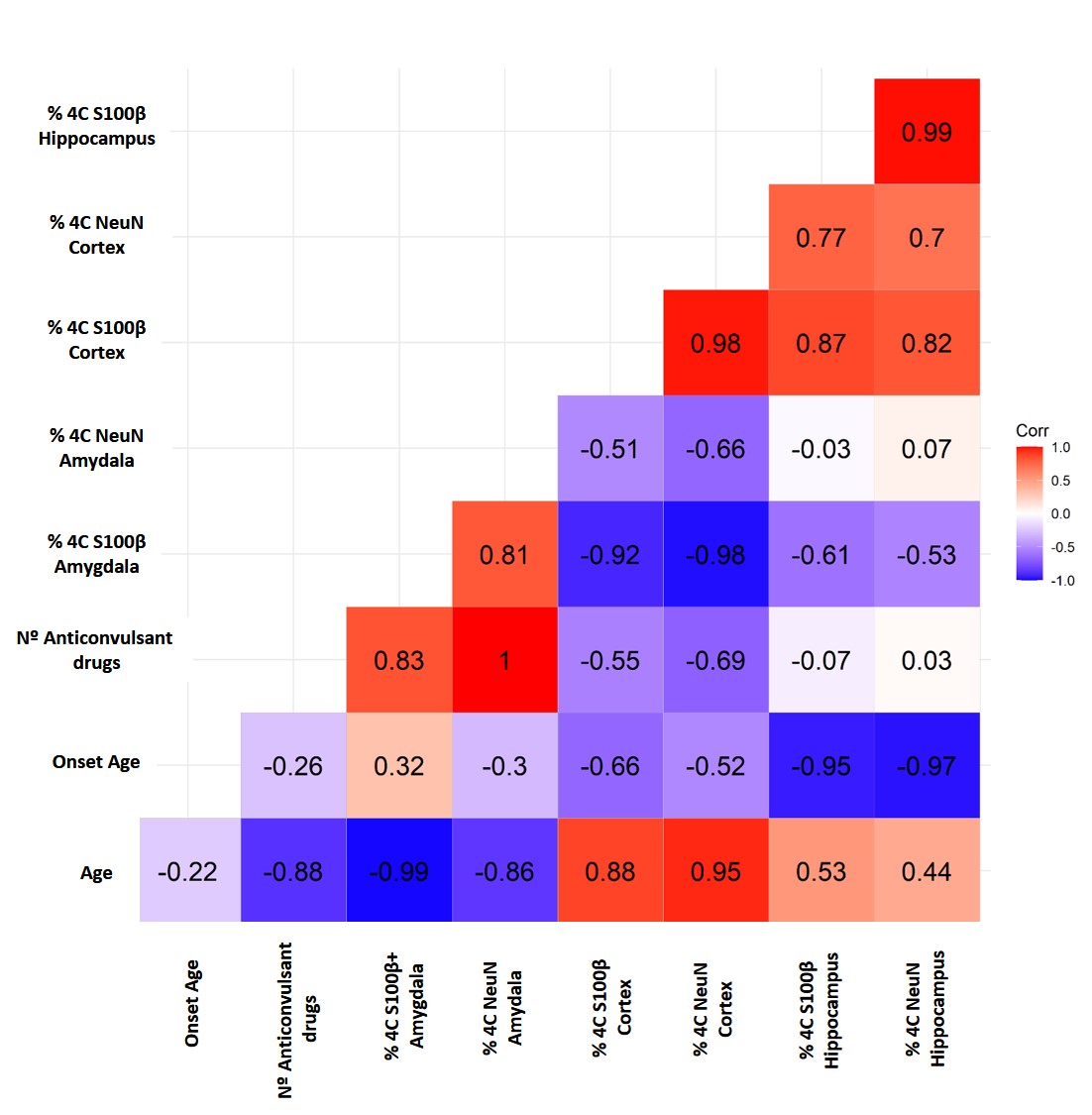
